# Adjunctive Ketamine for Sedation in Critically Ill Mechanically Ventilated Patients: An Active-Controlled, Pilot, Feasibility Clinical Trial

**DOI:** 10.1101/2021.04.26.21256072

**Authors:** Marwa Amer, Khalid Maghrabi, Mohammed Bawazeer, Kamel Alshaikh, Mohammad Shaban, Muhammad Rizwan, Rashid Amin, Edward De Vol, Mawadah Baali, Malak Altewerki, Mehreen Bano, Fawziah Alkhaldi, Sanaa Alenazi, Mohammed Hijazi

## Abstract

**Objective:** Ketamine has been shown to decrease sedative requirements in intensive care unit (ICU). Randomized trials are limited on patient-centered outcomes. We designed this pilot clinical trial to evaluate the feasibility of using ketamine as an adjunct analgosedative compared with standard of care (SOC) alone and determine preliminary effect size on 28-day mechanical ventilation (MV) duration and ventilator-free days (VFDs).

**Design:** Pilot, single-center, active-controlled, open-label, randomized clinical trial.

**Setting:** Medical, surgical, and transplant ICUs at King Faisal Specialist Hospital and Research Center, Saudi Arabia.

**Patients and Methods:** Adult patients who were intubated within 24 hours, expected to require MV for the next calendar day, and had institutional pain and sedation protocol initiated.

**Intervention:** Adjunct ketamine infusion 1-2 μg/kg/min for 48 hours versus SOC.

**Measurements and Main Results:** Total of 83 patients (43 in SOC and 40 in ketamine) were included. Demographics were balanced between groups. Median MV duration was 7 (interquartile range [IQR] 3-9.25 days) in ketamine and 5 (IQR 2-8 days) in SOC, p= 0.15. Median VFDs was 19 (IQR 0-24.75 days) in ketamine and 19 (IQR 0-24 days) in the SOC (p=0.70). More patients attained goal RASS score at 24 and 48 hours in ketamine (67.5% and 73.5%, respectively) compared with SOC (52.4% and 66.7%, respectively). Sedatives and vasopressors cumulative doses, and hemodynamic changes were similar. ICU length-of-stay was 12.5 (IQR 6-21.2 days) in ketamine, compared with 12 (IQR 5.5-23 days) in SOC, p=0.89. Consent and protocol adherence rate were adequate. No serious adverse events were observed in either group.

**Conclusions:** Use of ketamine as an adjunct analgosedative agent appeared to be feasible and safe with no negative impact on outcomes, including hemodynamics. The protocol of this pilot trial could be improved by modifying ketamine dosing regimen. These findings provide a basis for future, adequately powered, multicenter trial to investigate its association with patient-centered outcomes further.

## Background

Analgo-sedation or analgesia-first-sedation has gained popularity in recent years (1). This approach has been developed to decrease sedative use, and facilitate mechanical ventilation (MV) weaning (2). Data on ideal sedative in intensive care unit (ICU) for mechanically ventilated, and hemodynamically unstable patients are limited. Ketamine has a favorable hemodynamic, analgesic, and adverse effect profile, making it attractive as an analgosedative agent (3,4). It inhibits N-methyl-D-aspartate (NMDA) receptors and activates opioid μ- and κ-receptors (5). When compared with other sedatives, ketamine has fastest onset (within 30-40 sec) and 15 min duration of action. Anesthetists have long used ketamine for acute and chronic pain, procedural sedation, and rapid sequence intubation. It has also been used in postoperative pain control in surgical and trauma patients [as part of multimodal opioid-sparing analgesia in enhanced recovery after surgery (ERAS)], status asthmatics, status epilepticus, alcohol withdrawal, and agitation (6,7).

Ketamine does not appear to have potential side effects of nonsteroidal anti-inflammatory drugs or opioids negative effects on μ receptors of gastrointestinal tract associated with ileus (8-10). Studies to control acute pain in traumatic rib fractures of severely injured individuals at sub-anesthetic doses resulted in reduction of pain scale score and morphine-equivalent dose (11,12). Its use has been extended during coronavirus disease (COVID-19) pandemic due to a shortage of other sedatives to keep patients on MV comfortable and synchronous (13,14). Ketamine has not been associated with chest wall rigidity precipitating insufficient ventilation, which has occasionally been described with fentanyl [15]. Additionally, propofol and dexmedetomidine associated-hypotension may necessitate vasopressor support which may exclude patients from qualifying for COVID-19 antiviral medication (remdesivir), making ketamine an attractive alternative [16].

There is an increasing body of literature on use of ketamine infusion as an analgosedative agent at ICU to reduce sedative requirement and maintain patients within target sedation agitation scale goal (17-20). However, evidence provided in Pain, Agitation-Sedation, Delirium, Immobility, and Sleep Disruption (PADIS) guideline supporting its use in mechanically ventilated patients was insufficient due to limited number of randomized controlled trials (RCTs) (1,21). Data on whether ketamine affects patient-centered outcomes and its safety in RCTs, as compared with standard of care (SOC), are unclear. We evaluated the feasibility of an Analgo-sedative adjuncT keTAmine Infusion iN Mechanically vENTilated ICU patients (ATTAINMENT trial) compared to SOC alone, using a randomized trial design to determine preliminary estimates and effect size on patient-centered outcomes for future adequately powered clinical trial.

## Patients and Methods

This was an investigator-initiated, pilot, single-center, parallel-group, open-label RCT. The trial was registered with ClinicalTrials.gov: NCT04075006, current controlled trials: ISRCTN14730035, and Saudi Food and Drug Authority: SCTR #19063002. Our institution’s research ethics committee approved the trial (number 2191 187). Full study protocol has been published previously (22). The trial was conducted according to CONSORT guidelines extension to randomized pilot and feasibility trials. Participants were recruited from King Faisal Specialist Hospital and Research Center (KFSH&RC), Riyadh. It is a major referral center that provides tertiary and quaternary care. The ICU department is composed of several ICUs (medical, surgical, and transplant). During COVID-19 pandemic, new units were opened to accommodate patients surge.

### Study Population

Patients were eligible for inclusion if they were admitted to any of our three adult ICUs, intubated within previous 24-hours and expected to continue on MV the next calendar day, initiated on institutional pain and sedation protocol, and no objection from ICU attending or primary treating team. Recruitment began in September 2019 and was completed in November 2020. Patients were excluded if they had history of dementia or psychiatric disorders, or were comatose on admission due to hepatic encephalopathy. Other exclusion criteria were: severe pulmonary hypertension, tracheostomy at baseline, intellectual disability that precluded delirium assessment, transfer from an external facility, history of substance abuse, and situations where high blood pressure could trigger dangerous complications, such as aortic dissection. We also excluded those with repeated ICU admissions within same hospital visit and those who participated in another interventional trial. Full inclusion and exclusion criteria are detailed in **Supplementary Table 1**. Our research coordinators, along with local principal investigators screened patients for eligibility by using an electronic screening form in Research Electronic Data Capture (REDCap). Once eligibility criteria were met, informed consent was obtained. Given the need to enroll patients in expedited manner within 24-hours window, verbal consent from surrogate decision-maker (SDM) was allowed and documented in electronic medical records (EMR). Written consent was obtained as soon as SDM became available.

Patients were randomized in a 1:1 allocation using a computer-generated, pre-determined randomization list created by an independent biostatistician; no stratification was performed. Group allocation was concealed until after randomization. The investigators were masked to outcome data during the trial. Since this was an open-label study and because of lack of funding, blinding of investigators and treating team was not possible at this phase. However, patients and families were unaware of group assignment. **Supplementary Figure 1A, 1B** summarized treatment algorithm. After randomization, control group (SOC) was started on KFSH&RC ICU analgesia and sedation protocol. Since it was a nurse-driven protocol, treating team placed an order regarding target Richmond Agitation-Sedation Scale (RASS), and sedatives infusions were adjusted according to RASS target by bedside ICU nurse. For those randomized to intervention group (ketamine), continuous infusion ketamine 1-2 μg/kg/min was added as an adjunct for 48-hours. The infusion could be weaned off earlier in preparation for extubation. Since this was only a feasibility trial, there was no further intervention after 48-hours; however, clinical outcomes and adverse events (AEs) were monitored up to day 28. Ketamine dose was reported in μg per kilogram of actual body weight per min as per institutional practice. Other aspects of care, including fluid management, vasopressors use, blood products, enteral nutrition, renal replacement therapy, and early mobilization at discretion of treating team, were similar in both groups. ICU supportive measures were applied as appropriate, including venous thromboembolism prophylaxis, and prone positioning of patients who met the criteria based on established guidelines for acute respiratory distress syndrome (ARDS). Septic patients were managed according to latest survival sepsis campaign guidelines. Patient-ventilator asynchrony was systematically assessed and managed through inter-professional collaboration by prioritizing analgesia, and management of MV to avoid unnecessary use of neuromuscular blocking blockers [NMB]. Spontaneous awakening trial (SAT) was assessed every morning with SAT safety screen unless patients were receiving sedative infusion for status epilepticus or started on NMB post-randomization. Patients who passed SAT were immediately managed using spontaneous breathing trial (SBT) protocol. Both groups received basic analgesic regimen that included paracetamol and epidural analgesia for hyperthermic intraperitoneal chemotherapy (HIPEC) patients. If delirium treatment was needed, non-pharmacological measures (reassurance or mobilization, and family support) were applied first. If this was insufficient, the protocol allowed the use of antipsychotics and decision was left to ICU physician.

### Outcome Variables

Primary outcome was median duration of MV, with ventilator-free days (VFDs) up to day 28 as co-primary outcome. This outcome was chosen as patient-centered and highly influenced by mortality (23). Secondary outcomes included the following up to 28 days: ICU and hospital length-of-stay (LOS), mortality rate, and percentage of participants with AEs. We collected proportion and daily cumulative dose of vasopressors, sedatives and analgesics [fentanyl, propofol, midazolam, and dexmedetomidine], and antipsychotics over 48-hours post-randomization. Data on sedatives administered outside ICU settings during anesthesia or intraoperative management were not collected. Presence of delirium was assessed using confusion assessment method for ICU (CAM-ICU), which was measured at baseline and 48-hours post-randomization. If CAM-ICU scores were not available, an electronic progress note was reviewed to detect any evidence of delirium. Hemodynamic parameters [heart rate and mean arterial pressure (MAP)] were collected 48-hours post-randomization. Hemodynamic changes were defined as presence of tachycardia, hypertension, and hypotension. Details about variables collected and their definitions are available in **Supplementary File 1, and Supplementary Table 2**. Data were stored online in REDCap web application and data quality assessments were executed routinely to ensure completion and accuracy. Feasibility was assessed by evaluating consent rate, recruitment success, and protocol adherence. Consent rate was deemed to be adequate if > 70% of SDMs or patients chose to participate upon being approached. Successful recruitment was defined as > 3 patients enrolled per month. Protocol adherence was defined as >75% of patients receiving ketamine according to prescribed protocol. These thresholds were chosen after examining other pilot studies on complex interventions (22). We conducted educational sessions for clinicians, nurses, and hospital pharmacies to facilitate implementation of protocol. Protocol deviation was defined as not starting ketamine immediately after randomization (ideally within 4-hours) due to pharmacy delay or non-placement of ketamine order.

### Statistical analysis

Overall study population included all patients who were enrolled, randomly assigned, and received at least one dose of study medication, constituting modified intention-to-treat (mITT) population. Statistician was blinded to study group allocation and performed statistical analyses using R statistical software Version 3.5.0 (R Foundation for Statistical Computing, Vienna, Austria) and SAS/JMP version 15.0 (SAS Institute, Cary, NC). Counts and percentages were used to summarize distribution of categorical variables. Continuous variables were summarized using either mean ± standard deviation or median and interquartile range (IQR), based on results of normality testing (using Shapiro-Wilk test and Histograms). Chi-square test of independence was used to assess whether distribution of categorical variables was different between groups. Unpaired t-test or Mann-Whitney test was used to compare distribution of normal and non-normal continuous variables. Variables with more than one possible response were dummy coded, and percentage of each response was calculated from total sample size. VFDs were calculated by subtracting number of ventilation days from 28 after assigning VFD=0 for patients who died during 28 days. Sensitivity analysis for sedative and vasopressor requirements, excluding patients started on NBM post-randomization, was conducted. We ensured immediate data entry and identified missing data quickly, and issues were resolved promptly. Thus, no imputation for missing variables was done. Since this is a pilot and feasibility trial, there was no formal sample size calculation. A convenient sample of about 40 patients per group was thought to be roughly sufficient for this pilot trial (24).

## Results

From September 2019 through November 2020, a total of 437 MV patients were screened; 83 patients met inclusion criteria and 354 were excluded. Among screened patients, 88 did not meet eligibility criteria, mainly because they were expected to require MV for < 24-hours. Among included patients, 43 were in SOC group and 40 were included in ketamine group in mITT analysis. Participants’ flow through the trial is shown in **Figure 1**. Other exclusion criteria assessed were pulmonary hypertension (3 patients), tracheostomy at baseline post-face flab or due to subglottic stenosis (11 patients), intellectual disability that precluded delirium assessment (2 patients), transferred from outside facility (2 patients), and history of substance abuse (3 patients).

**Figure 1:**
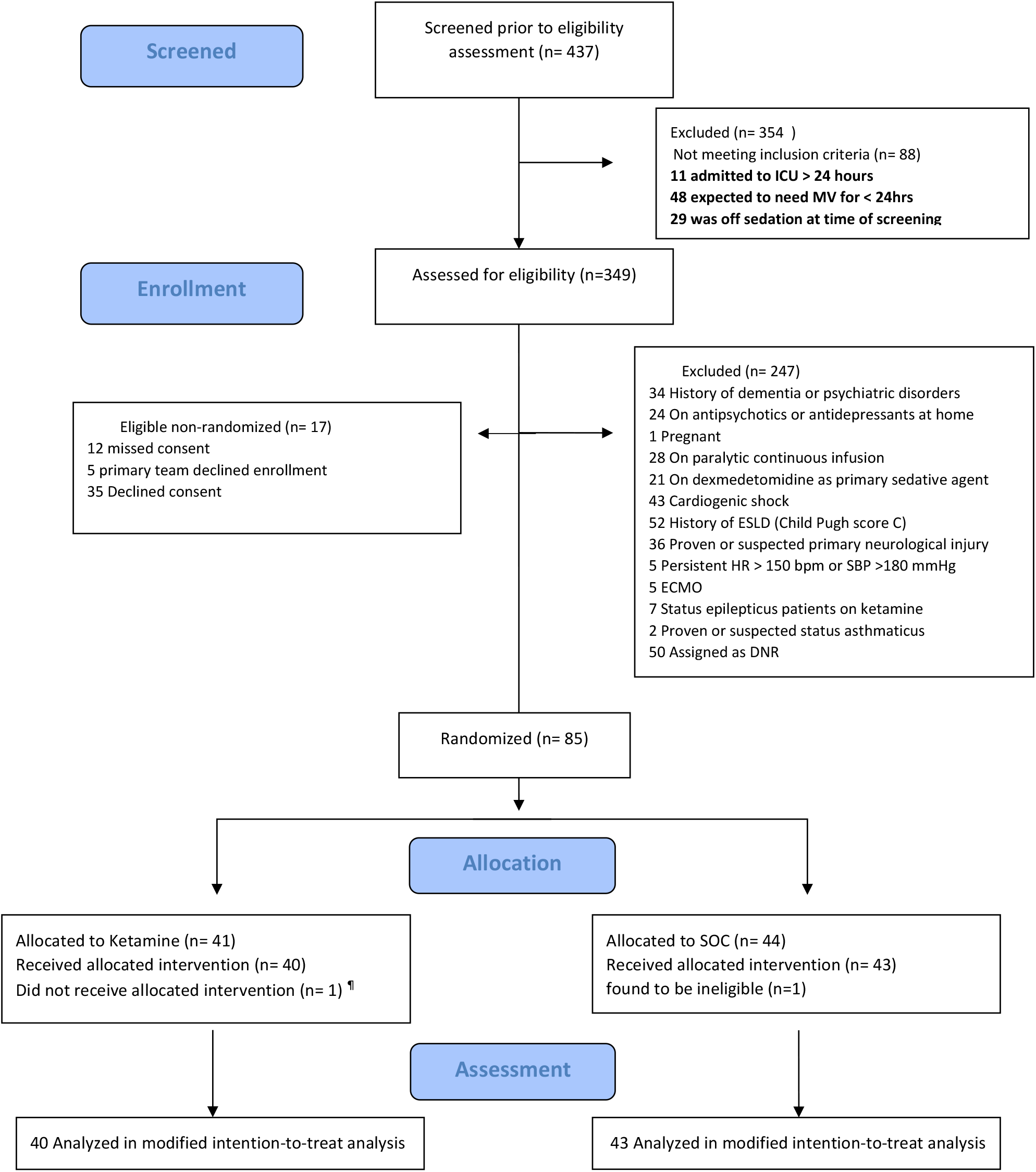
Study Flow Chart. ¶ deemed to be extubatable post-randomization Abbreviations: ESLD, end-stage liver diseases; HR, heart rate; SBP, systolic blood pressure; ECMO, extracorporeal membrane oxygenation; DNR, do-not-resuscitate

Baseline demographics are described in **Table 1 and Supplementary Table 3**. Median age was 60 years, with a higher proportion of males. About half of the patients were from medical ICU, 26.5% from surgical ICU, and 25.3% from transplant ICU; median SOFA 8, and APACHE II 20. Overall, demographic characteristics were balanced between groups in terms of sex, race, and comorbidities, except for prevalence of chronic obstructive pulmonary disease, which was higher in SOC group. We included a wide variety of ICU admission diagnoses and among those randomized to ketamine, 60% had ARDS and about 34% were recipients of solid organ transplants or had solid malignancy. Other primary reasons for ICU admission included HIPEC (3 patients: 2 in ketamine and 1 in SOC), COVID-19 pneumonia (2 patients: one in each group), and sickle cell disease (1 patient in ketamine). Ketamine-treated patients were noted to have higher median lactate level (2.2 [IQR 1.58-3.4 mmol/L] p= 0.004). Median number of hours of ICU admission before study enrollment was 13 (IQR 6-21.15) in SOC and 15 (IQR 12-21) in ketamine (p=0.17). Post randomization, NMB was initiated in 12.5% of ketamine-treated patients compared to 4.65% in SOC.

**Table 1.**
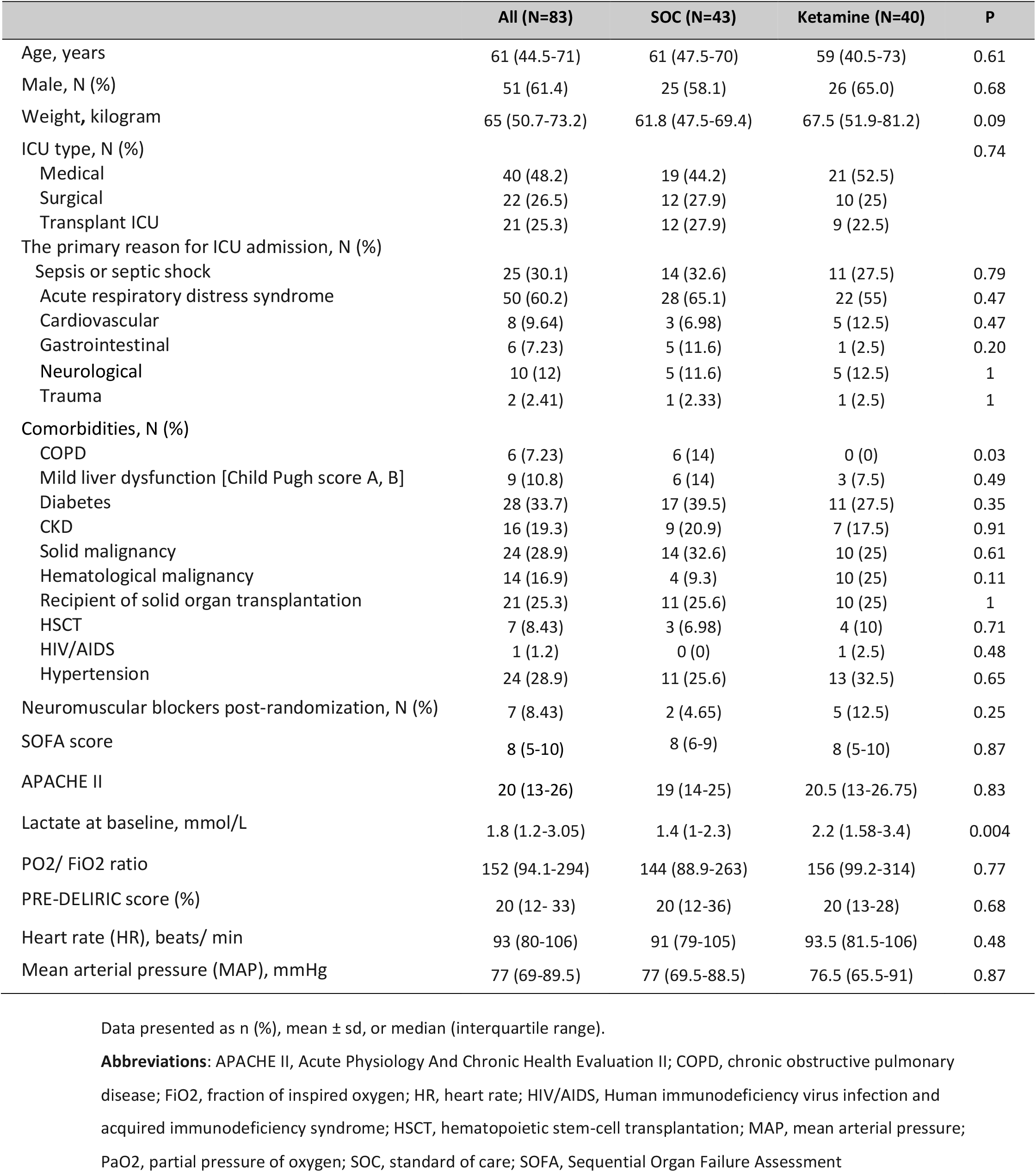
Demographic and baseline characteristics.

Patients outcomes are summarized in **Table 2**. Median duration of MV on day 28 was 7 days in ketamine group (IQR 3-9.25) compared to 5 days in SOC group (IQR 2-8). Among the 83 patients assessed, a similar proportion of patients had been weaned off MV at 28 days: 25 of 40 (62.5%) in ketamine group and 27 of 43 (62.8%) in SOC. Median distribution of VFDs at day 28 was 19 days in both groups (P = 0.70). Median duration of ICU LOS was comparable between groups. More patients in ketamine achieved goal RASS at 24 and 48 hours (67.5% and 73.5%, respectively) compared to SOC (52.4% and 66.7%, respectively). Additional details on outcomes are available in **Supplementary Table 4, Supplementary Figure 2**.

**Table 2.** Outcomes.

|  | All (N=83) | SOC (N=43) | Ketamine (N=40) | p |
| --- | --- | --- | --- | --- |
| **Primary and co-primary outcome** |  |  |  |  |
| MV within 28 days post-intubation, N (%) |  |  |  | 1 |
| No | 52 (62.7) | 27 (62.8) | 25 (62.5) |  |
| Yes | 31 (37.3) | 16 (37.2) | 15 (37.5) |  |
| 28-day duration of MV, days | 5 (2-9) | 5 (2-8) | 7 (3-9.25) | 0.15 |
| Duration of MV at ICU discharge/death, days | 8 (3-18.5) | 7 (3-13.8) | 9 (3-19) | 0.32 |
| Ventilation-free days, days <sup>a</sup> | 19 (0-24) | 19 (0-24) | 19 (0-24.75) | 0.70 |
| **Secondary** |  |  |  |  |
| Patients at goal RASS at 24 h, N (%)<sup>b</sup> | 49 (59.8) | 22 (52.4) | 27 (67.5) | 0.24 |
| Patients at goal RASS at 48 h, N (%)<sup>c</sup> | 51 (69.9) | 26 (66.7) | 25 (73.5) | 0.70 |
| Patients at Goal pain score at 24 h, N (%)<sup>d</sup> | 80 (96.39) | 41 (95.35) | 39 (97.5) | 1 |
| Patients at Goal pain score at 48 h, N (%)<sup>d</sup> | 79 (95.2) | 41 (95.3) | 38 (95) | 1 |
| Discharge from ICU, N (%) | 76 (91.6) | 40 (93) | 36 (90) | 0.71 |
| ICU length of stay, days | 12 (6-22.5) | 12 (5.5-23) | 12.5 (6-21.2) | 0.89 |
| Disposition at ICU discharge, N (%) |  |  |  | 0.64 |
| Morgue | 28 (33.7) | 16 (37.2) | 12 (30) |  |
| Floor | 55 (66.3) | 27 (62.8) | 28 (70) |  |
| Hospital discharge, N (%) | 79 (95.2) | 41 (95.3) | 38 (95) | 1 |
| Hospital length of stay, days | 26 (13-39) | 27 (12.5-47) | 26 (15.8-38) | 0.87 |
| Disposition at hospital discharge, N (%) |  |  |  | 0.96 |
| Home | 41 (49.4) | 21 (48.8) | 20 (50) |  |
| Morgue | 38 (45.8) | 20 (46.5) | 18 (45) |  |
| Another facility | 1 (1.2) | 1 (2.33) | 0 (0) |  |
| Still in hospital | 3 (3.61) | 1 (2.33) | 2 (5) |  |

Data presented as n (%), mean ± sd, or median (interquartile range).
<sup>a</sup> 53 patients (26 SOC and 27 ketamine) were alive and zero VFD was assigned for patients who died within 28 days
<sup>b</sup> The RASS measures levels of consciousness (scores range from -5 [unresponsive] to +4 [combative]). Assessed in 82 patients at 24 hours (42 SOC and 40 ketamine-treated)
<sup>c</sup> The RASS was assessed in 73 patients at 48 hours (39 SOC and 34 in ketamine-treated)
<sup>d</sup> Assessment of pain was done by Critical Care Pain Observation Tool for pain (CPOT)
**Abbreviations:** MV, mechanical ventilation; RASS, Richmond Agitation and Sedation; SOC, standard of care

Safety outcomes are described in **Table 3**. Proportion of patients who did not complete 48-hours of the trial was higher in ketamine (37.5%) than in SOC (11.63%) and the main reason was weaning off sedation in preparation for extubation. Antipsychotics were started in 3 ketamine-treated patients compared to 4 patients in SOC (p=1). Dexmedetomidine initiation within 48-hours post-randomization was similar between groups. Higher frequency of hypersalivation and frequent suctioning was observed in SOC arm. Regarding hemodynamic changes in HR and MAP at 24 and 48-hours, we found no difference between groups (**Figure 2**). The 28-day mortality rate was 11 (27.5%) in ketamine compared with 14 (32.6%) in SOC, p= 0.79. Data Safety Monitoring Committee reviewed all deaths, and all were determined to have been due to underlying disease, with participation in trial not being a contributing factor. More details of safety outcomes are available in **Supplementary Table 5**.

**Table 4.** Safety outcomes.

|  | All (N=83) | SOC (N=43) | Ketamine (N=40) | p |
| --- | --- | --- | --- | --- |
| 28-day Tracheotomy, N (%) | 22 (26.5) | 11 (25.6) | 11 (27.5) | 1 |
| 28-day unplanned extubation/Self-extubation, N (%) | 2 (2.41) | 0 (0) | 2 (5) | 0.23 |
| Patients who did not complete 48h of trial, N (%) | 20 (24.1) | 5 (11.63) | 15 (37.5) | 0.01 |
| Reason for trial discontinuation before 48h, N (%) |  |  |  |  |
| Excessive sedation and patients not in target RASS | 2 (10) | 0 (0) | 2 (13.3) | 1 |
| Death | 4 (20) | 2 (40) | 2 (13.3) | 0.25 |
| Goal changed to comfort care | 2 (10) | 1(20) | 1 (6.67) | 0.45 |
| Extubation and weaning off sedation in 48h | 14 (70) | 3 (60) | 11 (73.33) | 0.61 |
| Physician decline patient participation | 1 (5) | 0 (0) | 1 (6.67) | 1 |
| Hemodynamics |  |  |  |  |
| HR at 24h | 92 (75.5-107) | 95 (80-107) | 83.5 (71.8-105) | 0.11 |
| HR at 48h | 84 (72-100) | 89 (75 - 104) | 82 (71- 99) | 0.31 |
| MAP at 24h | 75 (64.5-87) | 75 (62.5 - 86.5) | 74.5 (69.5- 91.5) | 0.31 |
| MAP at 48h | 77 (65-90) | 76 (67.5-87) | 77.5 (64-92.5) | 0.50 |
| Uncontrolled agitation, N (%) | 10 (12.05) | 4 (9.3) | 6 (15) | 0.51 |
| Combative behavior to the nursing staff, N (%) | 2 (2.41) | 1 (2.33) | 1 (2.5) | 1 |
| Hyper-salivation and frequent suctioning, N (%) | 22 (26.5) | 14 (32.6) | 8 (20) | 0.29 |
| Patient started on antipsychotics within 48h post-randomization, N (%) | 7 (8.43) | 4 (9.3) | 3 (7.5) | 1 |
| Use of physical restraint 48h post-randomization, N (%) | 22 (26.5) | 10 (23.3) | 12 (30) | 0.66 |
| 28-day mortality rate, N (%) | 25 (30.1) | 14 (32.6) | 11 (27.5) | 0.79 |

Data presented as n (%), mean ± sd, or median (interquartile range).
**Abbreviations:** CAM-ICU, Confusion Assessment Method for the ICU; HR, heart rate; MAP, mean arterial pressure; RASS, Richmond Agitation and Sedation Scale; SOC, standard of care.

**Figure 2.**
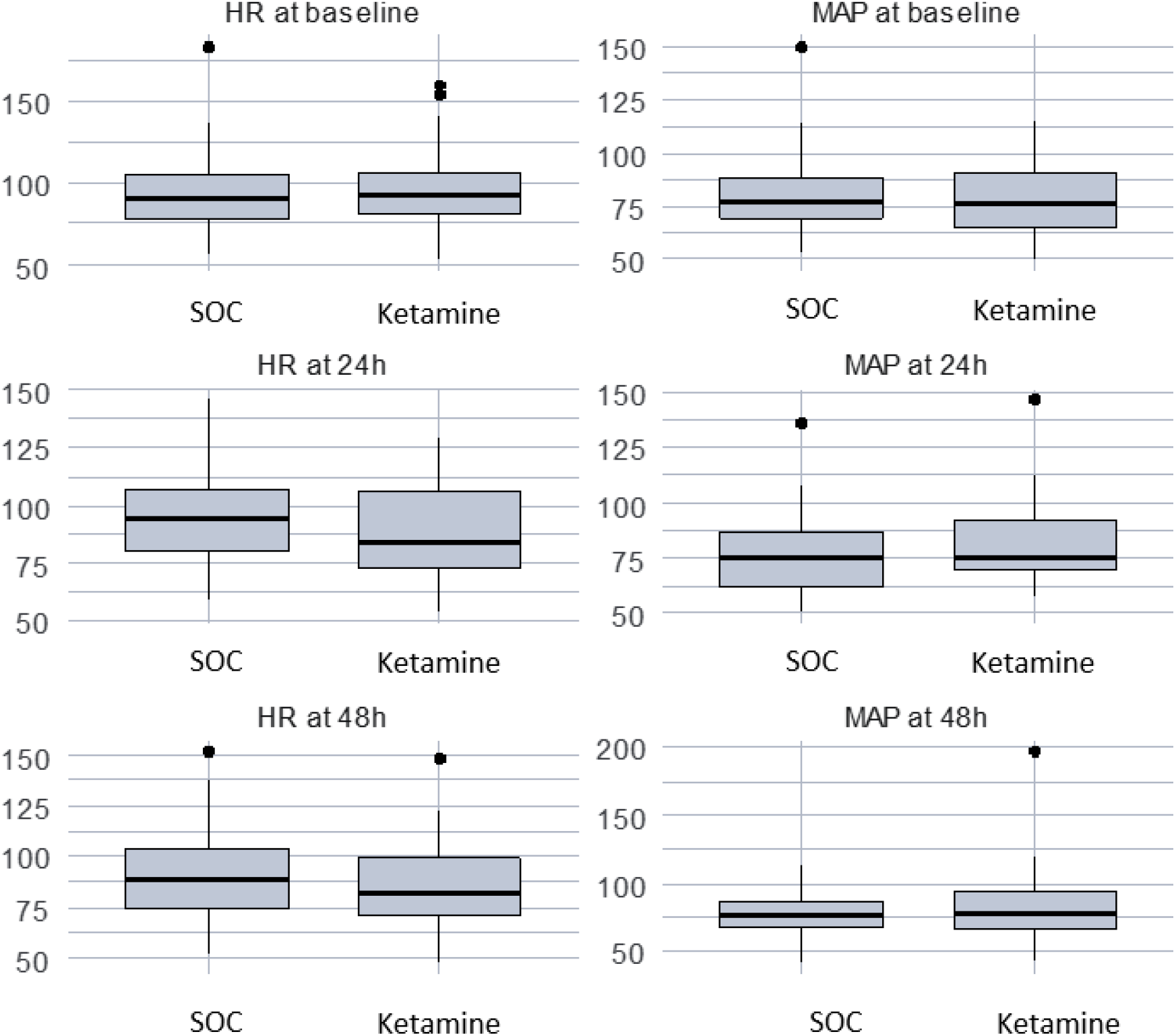
HR and MAP at baseline, 24 hours, and 48 hours. Abbreviation: SOC, standard of care; HR, heart rate; MAP, mean arterial pressure

**Supplementary Tables 6-7, Supplementary Figures 3-4** described sedative and vasopressor requirements. Median RASS was -2 at baseline, which gradually increased to -1 during 48-hours post-randomization, indicating light sedation and ability of patients to make eye contact with verbal stimulation. Thirty-six patients underwent CAM-ICU assessment within 48-hours post-randomization (43.37 %), of which 2 were positive in ketamine group (5%). There was no difference in baseline values of vasopressor and sedative requirements pre-randomization, except for amount of vasopressin which was higher in ketamine-treated patients (median 39.6, IQR 30.5-64.2 units, P = 0.053). Cumulative doses of fentanyl and other sedatives were similar between two groups at 48-hours post-randomization. Similar trends were observed for cumulative vasopressors dose in mg at 48-hours post-randomization. Sensitivity analysis was conducted on sedative requirements, excluding those started on NMB post-randomization, and findings were consistent with primary analysis.

Regarding feasibility outcomes, average patients enrollment was 3-4 patients/month. Consent rate was adequate; > 70% of SDMs or patients choosing to participate when approached for consent. Recruitment rate decreased significantly during COVID-19 pandemic and was halted for 1 month. We resumed recruitment at a slower rate in March 2020, with an average of 1-2 patients/month. In total, 12% of patients were enrolled outside traditional working hours (on weekends or night shifts). This process was facilitated through close collaboration with on-call ICU physician. Protocol adherence was 97.5% and median hours from consent or enrollment until ketamine started was 4.25 [IQR 2.08-5.88]. Adherence rate was lower than expected (90%) during COVID-19 pandemic. We were able to improve compliance using strategies such as education sessions for research and clinical staff and routine clinical reminders, including documentation in EMR.

## Discussion

This pilot proved the feasibility of a clinical trial to evaluate the use of ketamine as an analgosedative in ICU patients with MV. Achieving our threshold of recruitment and consent rate demonstrated that the trial is acceptable to clinicians, patients, and families. Major barriers faced were difficulties in continuing under lockdown conditions, infected research staff, shifting staff to cover COVID-19 ICU, and reorientation in clinical trial research towards COVID-19. We demonstrated that ketamine appeared to be safe, and majority of the patients achieved target RASS and pain scores. There was no increase in antipsychotics or dexmedetomidine use post-randomization and no notable hemodynamics changes. As this was a pilot study, it was not powered to detect statistically significant differences in patient-centered outcomes between the two groups.

Most previous studies had a limited focus on patient-centered outcomes as primary outcome favoring surrogate outcomes, such as sedation scores and changes in analgesics and sedatives, leaving a significant knowledge gap about the use of ketamine as a sedative agent in ICU. To the best of our knowledge, this pilot trial was the first that reported a patient-centered outcome as primary outcome and included diverse ICU population. We chose duration of MV as a primary outcome, because ketamine lowers airway resistance, preserves pharyngeal and laryngeal protective reflexes, increases lung compliance, and is less likely to cause respiratory depression in low and slow infusions (4). Median duration of MV and VFDs in our cohort was consistent with that reported in MENDS2 sedation trial; adjusted median, 23.7 days in dexmedetomidine vs. 24 days in propofol; OR, 0.96; 95% CI, 0.74-1.26) (25). It is noteworthy that majority of our population were from medical ICU (48.2% of entire cohort) and had moderate ARDS, with a median baseline PaO2/FiO2 ratio of 152. Patients with ARDS may be under-represented in analgesia/sedation studies and currently recommended ketamine dosing strategy in this pilot may not be feasible and could be improved by modifying the regimen in upcoming adequately powered, multicenter trial.

In this pilot trial, cumulative doses of fentanyl and other sedatives were similar between the two arms. This could be explained by the fact that proportion of patients who did not complete 48-hours of the trials was significantly different between groups and was higher in ketamine group (37.5%). This could also be due to starting NMB in five patients randomized to ketamine compared to two patients in SOC after randomization. In addition, protocol adherence rate and sedatives titration were lower than expected (90%) during COVID-19 pandemic, possibly due to re-assignment of ICU nurses to COVID-19 ICUs. Subsequently, newly hired non-ICU nurses were assigned to cover manpower shortages in non-COVID-19 ICUs and could be unaware of study protocol. Hence, efforts to reduce concomitant sedatives with ketamine may be conservative. A trial by Guillou *et al* which showed a reduction in opioid consumption with low-dose ketamine infusion for 48-hours (26). However, patients in this trial underwent postoperative abdominal surgery and were able to use patient-controlled analgesia. It is difficult to extrapolate these findings to mechanically ventilated patients who are unable to self-report pain and have a higher severity of illness as in our cohort.

Another question to address pertains to ketamine dosing for analgosedation. It is well known that severity of critical illness influences drug pharmacokinetics and pharmacodynamics (27). Hemodynamic instability, sympathetic overstimulation, and acute septic brain dysfunction negatively affect organ function and thus distribution, absorption, metabolism, and drug dose-response relationships. Severely ill patients need much lower doses of sedatives to maintain adequate sedation. Ketamine is highly lipophilic and is metabolized in liver, generating active compounds (norketamine and hydroxynorketamine), and is eliminated in urine with an elimination half-life of 1.5-3 hours (28,29). Published data for ketamine doses showed that it can be safely titrated up to 10-20 µg/kg/min, as needed, to achieve desired level of analgosedation. We chose ketamine dosing at 1-2 µg/kg/min, because majority of ICU population included in our pilot were older (median age 61 years), with renal and hepatic dysfunction, which potentially alters metabolism and excretion of ketamine and its active metabolite, resulting in increased sensitivity to ketamine, prolonged duration, drug accumulation, and possible longer recovery. Moreover, the dose described in this pilot was in agreement with existing literature describing light sedation strategy and 2018 PADIS guideline recommendations (1,13). This regimen is more conservative, to minimize dose-related reactions, such as psychotomimetic side effects, which could lead to complex differential diagnoses in ICU patients who are prone to delirium and other CNS disturbances. We also did not observe notable severe confusion, nightmares, emergence phenomena, or serious AEs associated with ketamine use, which is consistent with the findings reported by Perbert *et al* (30).

Ketamine has a sympathomimetic effect and can cause hypertension and tachycardia by acting as a catecholamine re-uptake inhibitor. However, in a subgroup of patients, particularly those with a catecholamine-depleted state, it can sometimes cause hemodynamic compromise and hypotension (31). It is recommended to be avoided in patients with a history of cardiac disease or hypertensive crisis, due to its myocardial depressant effect (32). In our pilot trial, we excluded patients with cardiogenic shock due to potential harm. Moreover, ketamine was not associated with clinically significant hemodynamic changes and appeared to be safe. There was no increase in vasopressor requirements post-randomization despite the fact that ketamine-treated patients were sicker at baseline, as evident by higher lactate level and higher vasopressin dose at baseline.

We noticed that 28-day mortality rate in our cohort was 30.1% (32.6% in SOC and 27.5% in ketamine), which was slightly higher than mortality rate reported in older sedation trials (MIDEX and PRODEX trial) (33). This was expected because we are a tertiary care hospital. APACHE II and SOFA scores also suggest that these data were derived from a cohort of critically ill patients. This is comparable to mortality rate in patients admitted to ICU with severe sepsis and shock and to all-cause mortality rate reported in more recent sedation trials, such as SPICE III trial (29.1% in both dexmedetomidine and usual-care groups) and MENDS2 trial (38% in dexmedetomidine group vs. 39% in propofol, HR, 1.06; 95% CI, 0.74 -1.52) (25,34,35).

This pilot trial had several strengths. Firstly, it included patient-centered outcomes, high rates of completed follow-up, and comprehensive assessments of AEs associated with ketamine use and its impact on hemodynamic response. We believe that our results provide incremental value in understanding the effects of ketamine. Adherence to mITT principle, randomization, and blinded outcome assessors limited potential sources of bias. Moreover, our trial included diverse ICU populations from medical, surgical, and transplant ICUs. We also made every effort to include eligible patients within a narrow randomization window (within 24-hours of intubation) to eliminate potential confounders with other co-interventions. Lastly, study protocol (design, study enrollment, and outcomes) aligns with the design of clinical trials evaluating sedation in critically ill adult MV patients (36).

We acknowledge limitations of our pilot trial. It was a single-center and did not include neurocritical care ICU patients, such as those with severe traumatic brain injury (TBI) and hydrocephalus. More recent systematic reviews of mixed acute brain populations (subarachnoid hemorrhage, tumors, and TBI) concluded that ketamine had no detrimental effect on intracranial pressure, ICU LOS, or mortality (37). Furthermore, we did not collect data on frequency and duration of prone positioning for ARDS patients who were made prone, or median change in PaO2/FiO2 ratio post-randomization, limiting the ability to determine the real benefit of ketamine in oxygenation post-randomization. Although we made efforts to validate the diagnosis of delirium and delirium assessment with CAM-ICU, we had a large proportion of patients (56.6 %) with un-assessed CAM-ICU, leaving a knowledge gap to be addressed in future trial. Finally, ketamine duration was limited to 48-hours due to the nature of this pilot trial and longer duration needs to be investigated in future. We believe that the trial protocol could be improved by modifying the current ketamine dosing regimen, capturing MV settings after randomization, and collecting data on other co-interventions (e.g. corticosteroids, prone positioning, and diuretics). Future trial may also consider looking at ketamine analgo-sedative effect in COVID-19 and neurocritical care ICUs. Finally, ketamine is not an expensive drug. Currently, no studies have evaluated cost-effectiveness of ketamine in management of sedation and analgesia and this should be considered in future.

## Conclusions

Ketamine is potentially attractive option for analgosedation. In our pilot trial, ketamine appeared to be safe, and feasible with no negative impact on outcomes, including hemodynamics. However, this pilot trial is not sufficiently powered to show a difference in clinical outcomes. While these data are encouraging, results generated from this pilot lay the foundation for a future adequately powered, multicenter trial to shed light on remaining questions and investigate the association with patient-centered outcomes further.

## Availability of data and material

The datasets used and analyzed during the current report are available from the corresponding author on reasonable request.

## Acknowledgments

We thank the ICU physicians, ICU nurses, ICU respiratory therapists, ICU physiotherapists, ICU satellite pharmacists, and ICU clinical pharmacists of KFSH&RC for their efforts to save the lives of ICU patients and their bravery in fighting against the COVID-19 pandemic. We also thank the Saudi Critical Care Trials Group for providing feedback on the study proposal, as well as participants and their families. Without their collective generosity, this trial would not have been possible.

## Supplementary Data

**Supplementary Table 1:** Full inclusion and exclusion criteria.

| **Inclusion criteria:** |
| --- |
| 1. Adults patients (>14 years)<br>2. Recently intubated and commenced on mechanical ventilation within the last 24 hours.<br>3. Admitted to one of the following ICUs (Medical, Surgical, transplant/oncology or COVID-19).<br>4. Expected to require MV longer than 24 hours.<br>5. Expected to be on the KFSH&RC sedation protocol.<br>6. There is no objection of the ICU attending for enrollment. |
| **Patients with the above inclusion criteria and has one of the following exclusion criteria were screened then excluded:** |
| • Patients with a history of dementia or psychiatric disorders or those on any antipsychotic or antidepressant medications at home.<br>• Pregnancy.<br>• Age < 14 years old.<br>• Expected to need MV < 24 h.<br>• Known hypersensitivity to ketamine.<br>• Patients with expected targeted RASS score of – 5, e.g., patients on continuous infusion neuromuscular blockade.<br>• Patients on dexmedetomidine as the primary sedative prior to randomization.<br>• Patients with cardiogenic shock, acute decompensated heart failure, or myocardial infarction<br>• History of end-stage liver failure (Child-Pugh score C).<br>• Proven or suspected primary neurological injury (traumatic brain injury, ischemic stroke, intracranial hemorrhage, spinal cord injury, anoxic brain injury, brain edema).<br>• Patients with persistent heart rate (HR) > 150 beats per minute (bpm) or systolic blood pressure (SBP) >180mmHg.<br>• Patients identified as Do Not Resuscitate (DNR) or those expected to die within 24 h.<br>• Patients on extracorporeal membrane oxygenation (ECMO).<br>• Patients with refractory status epilepticus who are receiving ketamine infusion.<br>• Proven or suspected status asthmaticus (the dose of this indication differed from the recommended dose for analgosedation) |

**Supplementary Figure 1A:**
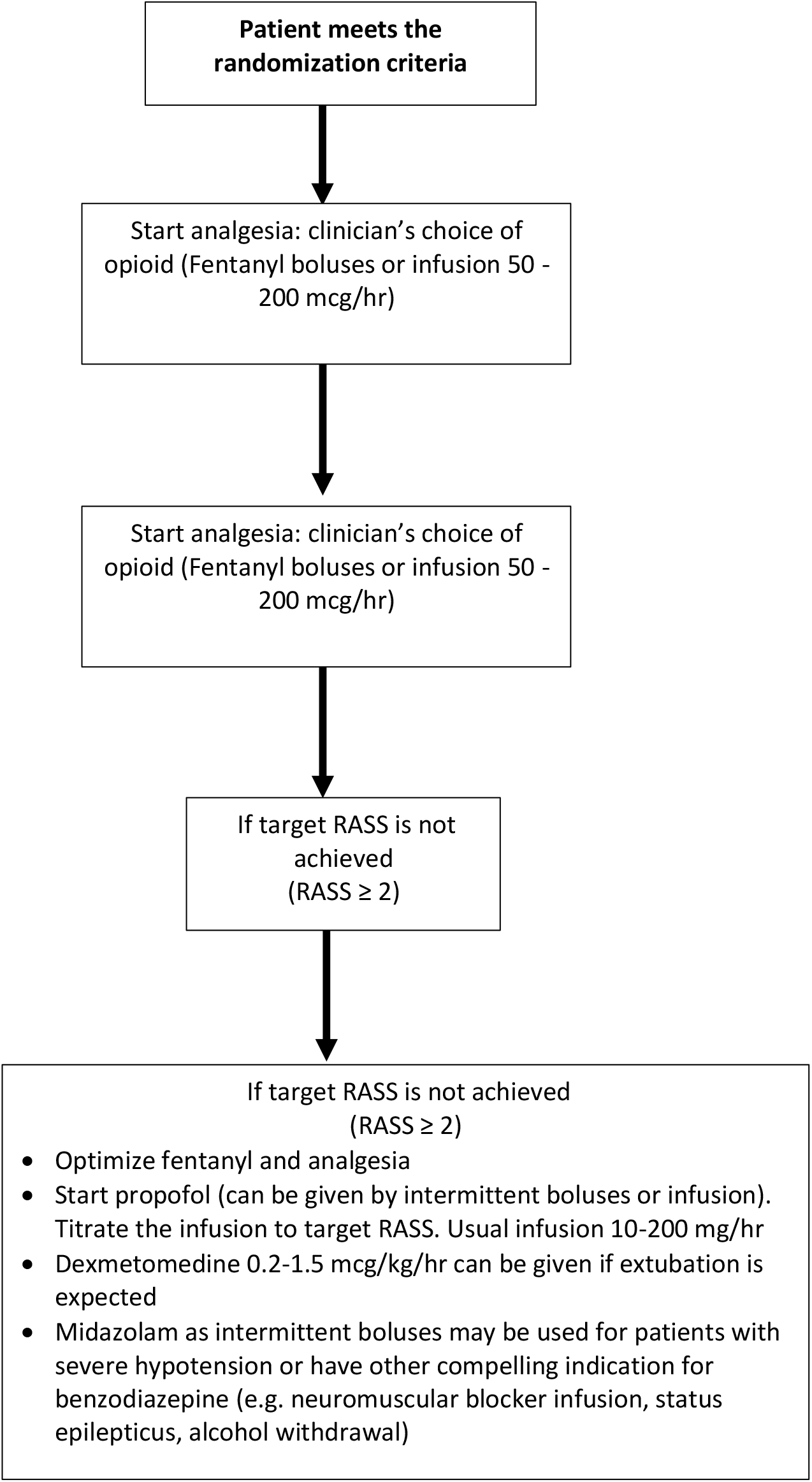
Treatment algorithm for Patient Randomized to Standard of care.

**Supplementary Figure 1B:**
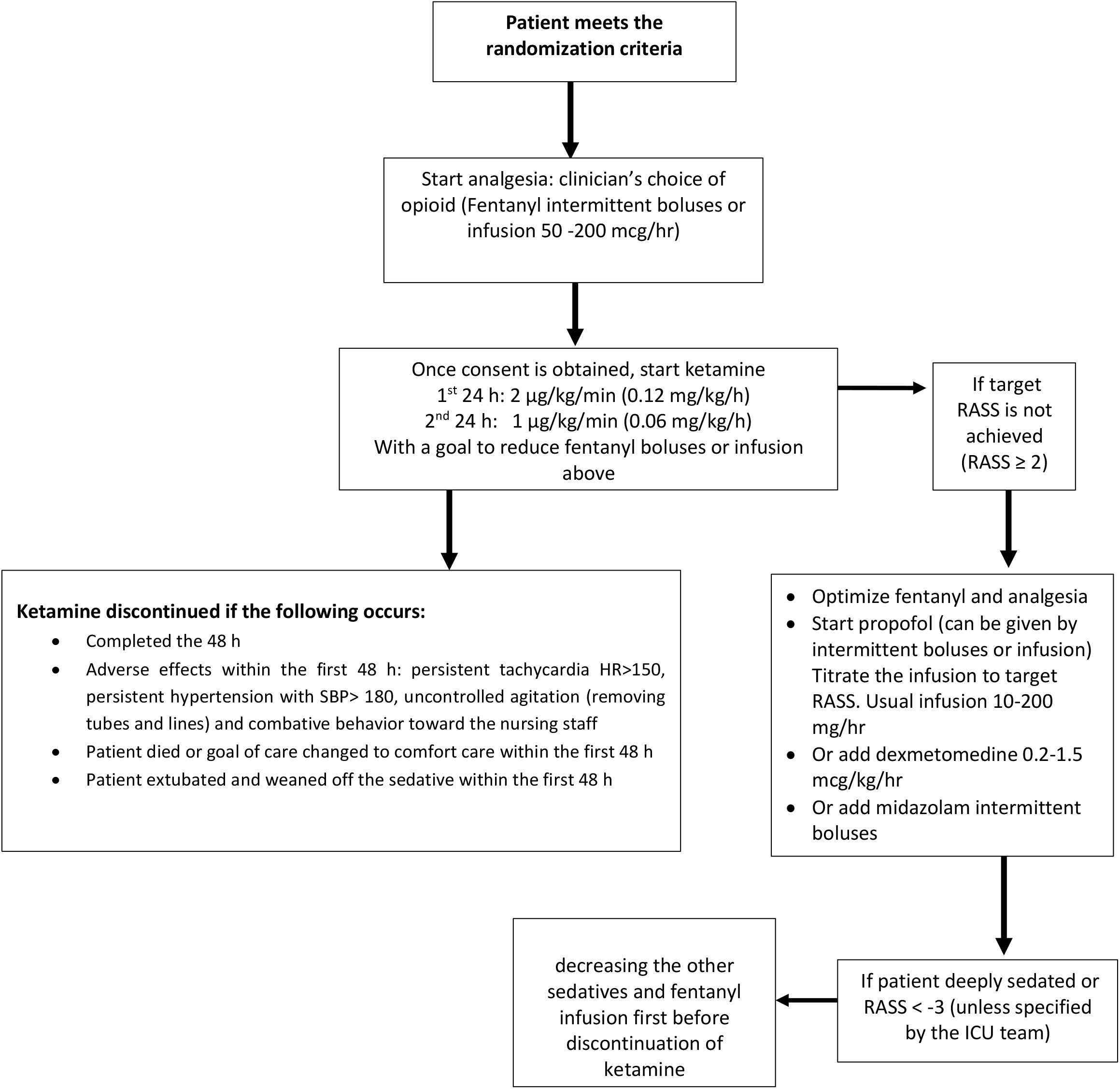
Treatment algorithm for Patient Randomized to Ketamine.

**Supplementary Table 2:** Details of outcome variables definition.

|  |  |
| --- | --- |
| Severity of illness | • Estimated by Sequential Organ Failure Assessment (SOFA) score and Acute Physiology and Chronic Health Evaluation (APACHE II) score, with higher scores indicating higher severity of illness<br>• The Sequential Organ Failure Assessment (SOFA) is used to track organ failure in the ICU; scores range from 0 to 24, with higher scores indicating greater severity of illness<br>• The Acute Physiology and Chronic Health Evaluation (APACHE II) assesses the risk of death on a scale from 0 to 71, with higher scores indicating a higher risk of death. |
| PRE-DELIRIC<br>Delirium Risk Score | • A delirium prediction model specifically designed for adult critical care patients 24 h after ICU admission. This model was used to predict the factors that may influence delirium risk prior to randomization |
| Duration of MV | • Was recorded as either the number of calendar days from intubation to extubation or until ICU discharge or death, whichever occurred first.<br>• This outcome was chosen as a patient-centered outcome and based on the mechanistic plausibility data that showed ketamine possibly has a bronchodilatory effect and maintains respiratory drive and airway reflexes |
| Successful extubation | • Was defined as the absence of the need for reintubation within 48 hours. |
| Cumulative dose of pain and sedatives | • Reported as proportion and median dose in the first 48 h after randomization |
| Patients achieving the RASS goal and pain score goal | • Reported as proportion of patients achieving this goal within the first 24 and 48 h after Randomization<br>• The RASS is a scale used to assess the depth of sedation on a scale of – 5 to + 4, with a negative value indicating deeper sedation and positive values indicating increased agitation<br>• Assessment of pain was done by Critical Care Pain Observation Tool for pain (CPOT) by evaluating facial expression, body movement, muscle tension, and adherence to use of the ventilator if intubated or vocalization if extubated. Total scores range from 0 to 8, with scores higher than 2 indicating the presence of pain. |
| vasopressor requirements | • Reported as proportion and median vasopressor requirements in the first 48 h after randomization |
| ICU and hospital LOS | • Number of calendar days (median, IQR) from randomization to discharge date from the ICU or hospital |
| Mortality rate | • Reported as proportion at the time of hospital discharge or 28 days after randomization, whichever comes first |
| Adverse events | • Any clinically significant worsening in a study participant's condition based on clinical judgment compared to the baseline status at the time of randomization will be recorded as an AE. This is applied whether or not the AE is considered to be related to the study treatment.<br>• Tachycardia was defined as a heart rate > 130 beats per minute<br>• Hyper or hypotension was classified as systolic blood pressure $\geq 180$ mmHg and $\leq 90$ mmHg, respectively<br>• Tracheostomy, unplanned extubation (self-extubation), and re-intubation Reported as proportion of patients within 28 days post-randomization<br>• Hypersalivation: defined as frequent suctioning the first 48 h after randomization (defined as interval between suctioning episodes 2 h or less). We calculated the modified Clinical Pulmonary Infection Score (CPIS) to differentiate secretions caused by patients' underlying lung pathology (ventilator-associated pneumonia [VAP]) vs ketamine-associated hypersalivation<br>• Physical restraint was reported as proportion of patients within 48 h after randomization<br>• The incidence of delirium was reported as proportion of patients starting on antipsychotics and positive CAM-ICU score to assess the incidence of delirium 48 h after randomization. The presence of delirium will also be confirmed through a psychiatrist consultation<br>• We believe the administration of sedative agents is standard of practice in the ICU to minimize a patient's discomfort while on MV. Hence, the expected adverse effects will not exceed what is encountered during daily practice (e.g., benzodiazepine-associated delirium, opioid-induced constipation, hemodynamic instability associated with propofol and dexmedetomidine, ketamine-associated sympathetic stimulation "tachycardia and increase in blood pressure," and possible delirium). |
| Serious adverse events | • Included death or potentially life-threatening adverse effects that requires inpatient hospitalization or prolongation of hospitalization, or results in permanent or significant disability/incapacity, congenital anomaly/birth defect, or the investigator considers significant.<br>• An independent Research Advisory Council at our institution served as a Data Safety Monitoring Committee (DSMC) and reviewed all adverse events (including all deaths). This committee included faculty with expertise in various disciplines engaged in human subjects' research from the hospital and research centre, and also community members. Consultants with special expertise might be invited to assist from time to time with complex issues. The committee will undertake periodic reviews at the discretion of the Chair, and an expedited review is done for all serious unexpected adverse events (SUAEs). The committee has the authority to suspend or halt recruitment if necessary.<br>• No formal interim analysis of efficacy will be undertaken due to possible small numbers that might preclude determination of a statistically significant difference in outcomes between the arms. No stopping rules is specified. |

**Supplementary Table 3.** Other demographic and baseline characteristics.

|  | All (N=83) | SOC (N=43) | Ketamine (N=40) | P |
| --- | --- | --- | --- | --- |
| Other primary reason for ICU admission, N (%) |  |  |  |  |
| Metabolic and endocrine disorder | 1 (1.2) | 1 (2.33) | 0 (0) | 1 |
| Renal | 6 (7.23) | 3 (6.98) | 3 (7.5) | 1 |
| Hematological | 3 (3.61) | 1 (2.33) | 2 (5) | 0.61 |
| Major surgery | 25 (30.1) | 11 (25.6) | 14 (35) | 0.49 |
| Source of infection, N (%) |  |  |  |  |
| Gastrointestinal | 5 (6.02) | 2 (4.65) | 3 (7.5) | 0.67 |
| Urine | 6 (7.23) | 4 (9.3) | 2 (5) | 0.68 |
| Blood | 9 (10.8) | 3 (6.98) | 6 (15) | 0.30 |
| Skin and soft tissue infections and osteomyelitis | 2 (2.41) | 2 (4.65) | 0 (0) | 0.49 |
| Respiratory | 26 (31.3) | 13 (30.2) | 13 (32.5) | 1 |
| Unknown | 11 (13.3) | 4 (9.3) | 7 (17.5) | 0.44 |
| Renal replacement therapy at baseline, N (%) | 7 (8.43) | 3 (6.98) | 4 (10) | 0.71 |
| Type of RRT, N (%) |  |  |  | 1 |
| iHD | 5 (71.4) | 2 (66.7) | 3 (75) |  |
| CVVH | 1 (14.3) | 1 (33.3) | 0 (0) |  |
| CVVHDF | 1 (14.3) | 0 (0) | 1 (25) |  |
| Urea at baseline, mmol/L | 7.2 (4.25-14.4) | 7.7 (4.65-17.6) | 6.15 (4.12-10.9) | 0.27 |
| Mode of mechanical ventilation, N (%) |  |  |  | 0.08 |
| AC | 43 (51.8) | 19 (44.2) | 24 (60) |  |
| PCV | 19 (22.9) | 14 (32.6) | 5 (12.5) |  |
| PS | 1 (1.2) | 1 (2.33) | 0 (0) |  |
| SIMV | 1 (1.20) | 1 (2.33) | 0 (0) |  |
| PRVC | 19 (22.9) | 8 (18.6) | 11 (27.5) |  |
| PH from ABG | 7.31 (7.24-7.40) | 7.34 (7.25-7.42) | 7.3 (7.23-7.37) | 0.31 |
| PCO2 from ABG, kilopascals | 6 (5-7.5) | 6 (5.1-7.6) | 5.9 (4.97-7.34) | 0.56 |
| PO2 from ABG, kilopascals | 10.5 (7.45-14.1) | 10.6 (6.85-14.1) | 10.1 (8.15-14.2) | 0.79 |
| HCO3 from ABG, mmol/L | 23.6 (20.1-29.2) | 23.7 (20.4-31.3) | 23.3 (19.5-27.5) | 0.31 |
| Metabolic acidosis, N (%) | 29 (34.9) | 15 (34.9) | 14 (35) | 1 |
| FiO2 | 0.5 (0.35-0.7) | 0.5 (0.4-0.7) | 0.5 (0.34-0.82) | 0.66 |

**Abbreviations:** AC, assist-control mode; ABG, arterial blood gas; CKD, chronic kidney disease; CVVH, continuous veno-venous hemofiltration; CVVHDF, Continuous veno-venous hemodiafiltration; FiO2, fraction of inspired oxygen; IHD, intermittent hemodialysis; RRT, renal replacement therapy; PaO2, partial pressure of oxygen; PCO2, partial pressure of carbon dioxide; PCV, pressure control ventilation; PS, pressure support; PRVC, pressure-regulated volume control; SIMV, Synchronized intermittent mandatory ventilation

**Supplementary Figure 2:**
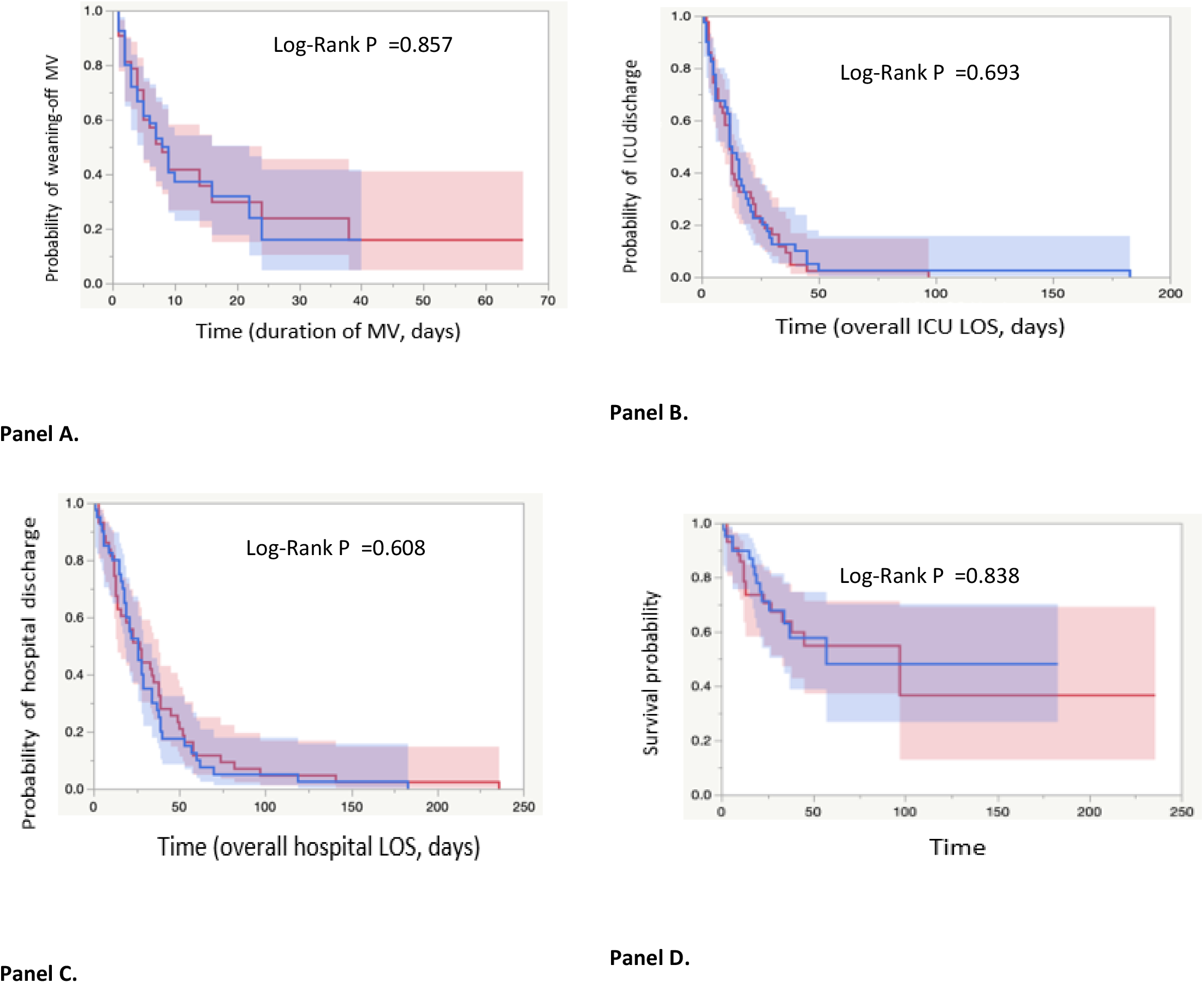
Time-to-event and Kaplan-Maier Curves. Kaplan-Meier curve with a corresponding log-rank test was used to estimate the probability of weaning-off MV. Patients who were not weaned-off MV during their ICU stay were censored at the last follow-up date. Kaplan-Maier estimates with a corresponding log-rank test were also used to compare the survival probability, hospital and ICU LOS between patients who received ketamine and patients who did not. Patients who alive or stayed in the ICU were censored at the last follow-up date. Panel A for duration of mechanical ventilation; Panel B represents the overall ICU length of stay (days); Panel C represents the overall hospital length of stay (days); Panel D represents 28-day mortality. Red line represents the standard of care group and blue line represents the ketamine group. Confidence interval was illustrated as a band around the time-to-event curves. SOC donates to standard of care

**Supplementary Table 4:** Cox-proportional regression analysis for weaning off mechanical ventilation.

| *Predictors* | *HR* | *95% CI* | *p* |
| --- | --- | --- | --- |
| Age | 0.99 | 0.97 – 1.01 | 0.352 |
| APACHE II on admission | 0.99 | 0.94 – 1.03 | 0.620 |
| SOFA score | 0.97 | 0.87 – 1.08 | 0.536 |
| Urea at baseline (mmol/L) | 1.02 | 0.99 – 1.04 | 0.235 |
| ketamine: No | *Reference* |  |  |
| ketamine: Yes | 1.19 | 0.66 – 2.16 | 0.563 |
| Metabolic acidosis: No | *Reference* |  |  |
| Metabolic acidosis: Yes | **0.48** | 0.24 – 0.94 | **0.032** |
| ICU type: Medical | *Reference* |  |  |
| ICU type: Surgical | **2.09** | 1.06 – 4.14 | **0.034** |
| ICU type: transplant | **2.11** | 1.02 – 4.35 | **0.043** |

**Table legend:** Abbreviation: APACHE II, Acute Physiology and Chronic Health Evaluation II; SOFA,
Sequential Organ Failure Assessment; HR : hazard ratio; CI: confidence interval
Cox-proportional regression analysis was used to assess factors associated with probability for weaning off MV and was reported as hazard ratio (HR) and 95% confidence interval (CI). The adjustment of observed effects was undertaken using a list of a priori defined covariates [age, Acute Physiology And Chronic Health Evaluation II (APACHE-II) score, Sequential Organ-Failure Assessment (SOFA) score, comorbidities, and ICU type].
The Cox-proportional hazard regression model showed that a lower probability of weaning off MV in patients with metabolic acidosis (HR [95% CI]): 0.48, [0.24-0.94], $P = 0.032$ ), whereas ICU admission for surgical reasons and transplant or oncology reasons was associated with a higher probability of weaning off MV [HR (95% CI): 2.09 (1.06–4.14) for surgical ICU, 2.11 (1.02–4.35) for transplant ICU] compared to medical admission

**Supplementary Table 5:**
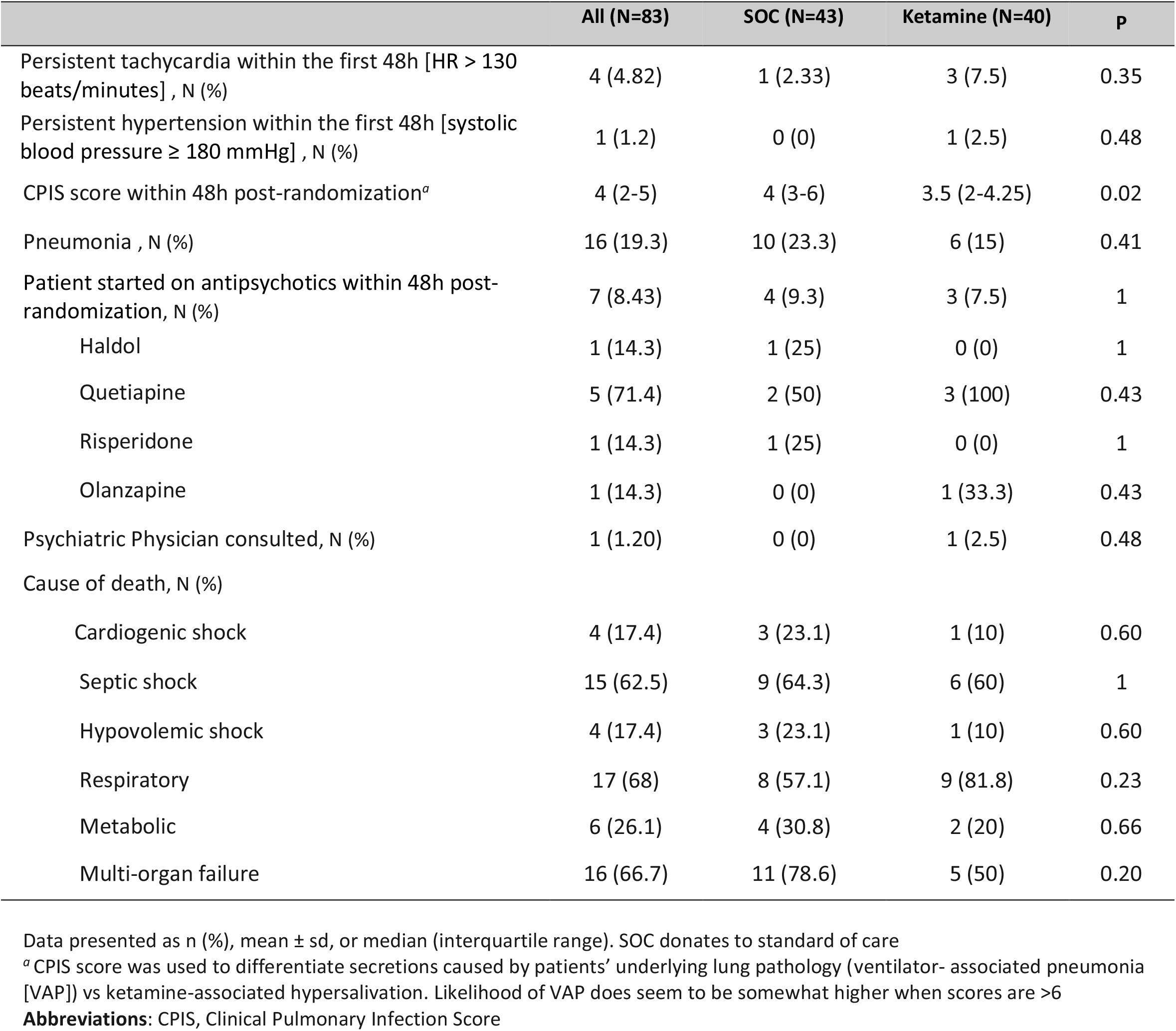
Other safety outcomes.

**Supplementary Table 6:** Sedatives and vasopressors requirements.

|  | Baseline |  |  |  | 48h post-randomization |  |  |  |
| --- | --- | --- | --- | --- | --- | --- | --- | --- |
|  | All (N=83) | SOC (N=43) | Ketamine (N=40) | P | All (N=83) | SOC (N=43) | Ketamine (N=40) | P |
| RASS Score <sup>a</sup> | -2 (-4 to -1) | -2 (-4 to -1) | -2.50 (-4 to -0.25) | 0.76 | -1 (-3 to -1) | -1.50 (-3 to -1) | -1.00 (-3 to 0) | 0.85 |
| pain score <sup>b</sup> | 0 (0-0) | 0 (0-0) | 0 (0-0) | 0.60 | 0 (0-0) | 0 (0-0) | 0 (0-0) | 0.31 |
| CAM-ICU Positive, N (%) <sup>c</sup> | 1 (1.2) | 0 (0) | 1 (2.5) | 0.58 | 2 (2.41) | 0 (0) | 2 (5) | 0.30 |
| CAM-ICU Negative, N (%)<sup>c</sup> | 36 (43.37) | 19 (44.19) | 17 (42.5) |  | 34 (40.96) | 19 (44.19) | 15 (37.5) |  |
| CAM-ICU Not assessed, N (%)<sup>c</sup> | 46 (55.42) | 24 (55.81) | 22 (55) |  | 47 (56.63) | 24 (55.81) | 23 (57.5) |  |
| Fentanyl, N (%) | 80 (96.4) | 40 (93.0) | 40 (100) | 0.24 | 82 (98.8) | 43 (100) | 39 (97.5) | 0.48 |
| Propofol, N (%) | 70 (84.3) | 35 (81.4) | 35 (87.5) | 0.64 | 48 (57.8) | 24 (55.8) | 24 (60) | 0.87 |
| Midazolam, N (%) | 47 (56.6) | 24 (55.8) | 23 (57.5) | 1 | 14 (16.9) | 8 (18.6) | 6 (15) | 0.89 |
| Dexmedetomidine, N (%) | . | . | . |  | 16 (19.3) | 12 (27.9) | 4 (10) | 0.05 |
| Cumulative dose of fentanyl (µg) | 1475 (681-2600) | 1262 (488-2612) | 1612 (1100-2512) | 0.17 | 3938 (2100-6400) | 3817 (2220-6140) | 4400 (1588-7700) | 0.67 |
| Fentanyl daily dose (µg /Kg) | 23.4 (9.9 - 39.7) | 21 (7.37-37.2) | 26.4 (15.7-43.2) | 0.22 | 66.8 (26.6-105) | 63.5 (32.8-97.1) | 69.6 (22.7-110) | 0.69 |
| Cumulative dose of propofol (mg) | 755 (172-1738) | 780 (150-1425) | 640 (215-1850) | 0.37 | 1990 (530-3862) | 2091 (492-3316) | 1815 (778-4272) | 0.95 |
| Propofol daily dose (mg/Kg) | 10.9 (3.26-24.7) | 12.7 (2.13-22.2) | 10.6 (4.38-25.3) | 0.63 | 28.4 (9.29-59) | 28.4 (6.59-58.1) | 28 (9.62-60.9) | 1 |
| Cumulative dose midazolam (mg) | 5 (3 -5.75) | 4.75 (2-5.38) | 5 (3 - 6) | 0.54 | 12.5 (5.25-101) | 7 (4.5 -32.2) | 62.8 (17.1-125) | 0.11 |
| Midazolam daily dose (mg/Kg) | 0.08 (0.04-0.13) | 0.08 (0.04-0.13) | 0.08 (0.05-0.12) | 0.89 | 0.24 (0.1 -1.25) | 0.15 (0.08-0.49) | 0.85 (0.26-1.74) | 0.16 |
|  | All (N=83) | SOC (N=43) | Ketamine (N=40) | P | All (N=83) | SOC (N=43) | Ketamine (N=40) | P |
| Cumulative Dexmedetomidine dose (µg) | . | . | . |  | 667 (357-1222) | 667 (357-1222) | 711 (310-1730) | 0.90 |
| Dexmedetomidine Daily dose (µg /Kg) | . | . | . |  | 9.34 (5.33-22.8) | 9.34 (5.33-22) | 18 (4.67-35.5) | 0.63 |
| Norepinephrine, N (%) | 50 (60.2) | 26 (60.5) | 24 (60) | 1 | 52 (62.7) | 27 (62.8) | 25 (62.5) | 1 |
| Epinephrine, N (%) | 6 (7.23) | 4 (9.3) | 2 (5) | 0.68 | 5 (6.02) | 1 (2.33) | 4 (10) | 0.19 |
| Phenylephrine, N (%) | 24 (28.9) | 16 (37.2) | 8 (20) | 0.14 | 11 (13.3) | 5 (11.6) | 6 (15) | 0.89 |
| Vasopressin, N (%) | 8 (9.64) | 5 (11.6) | 3 (7.5) | 0.71 | 15 (18.1) | 8 (18.6) | 7 (17.5) | 1 |
| Dopamine, N (%) | 4 (4.82) | 2 (4.65) | 2 (5) | 1 | 5 (6.02) | 3 (6.98) | 2 (5) | 1 |
| Cumulative dose of Norepinephrine (mg) | 5.92 (2.5-12.1) | 5.92 (1.82-10.7) | 6.35 (3.53-14.2) | 0.38 | 9 (4.92-28) | 8.63 (6.13-26) | 9.37 (4.4-28.4) | 0.89 |
| Cumulative dose of epinephrine (mg) | 1.17 (0.43-1.37) | 1.24 (0.53-1.5) | 0.81 (0.47-1.15) | 0.36 | 6.09 (2-13.1) | 29.2 (29.2-29.2) | 4.04 (1.88-7.84) | 0.16 |
| Cumulative phenylephrine dose (mg) | 0.45 (0.3-1.1) | 0.45 (0.3-1.3) | 0.45 (0.3-0.7) | 0.78 | 0.60 (0.21- 46.8) | 36 (0.5-57.6) | 0.45 (0.17-5.1) | 0.52 |
| Cumulative dose of dopamine (mg) | 133 (70.6 -203) | 133 (110-156) | 149 (86.2- 213) | 1 | 563 (490-676) | 563 (482-619) | 602 (546-659) | 0.56 |
| Cumulative vasopressin (units) | 18.2 (11.4-27.9) | 12 (9.6-15) | 39.6 (30.5-64.2) | 0.05 | 70.8 (30-91.6) | 24 (21.6 - 82.8) | 81.6 (60 - 89.6) | 0.20 |

Data presented as n (%), mean ± sd, or median (interquartile range). SOC donates to standard of care
<sup>a</sup> The RASS measures levels of consciousness (scores range from −5 [unresponsive] to +4 [combative])
<sup>b</sup> Assessment of pain was done by Critical Care Pain Observation Tool for pain (CPOT)
<sup>c</sup> The CAM-ICU, scores delirium as either present [positive] or not present [negative]]. Assessments were done when the patient was maximally awake. if in coma, unable to evaluate.
**Abbreviation:** CAM-ICU, Confusion Assessment Method for the ICU; RASS, Richmond Agitation and Sedation Scale.

**Supplementary Figure 3:**
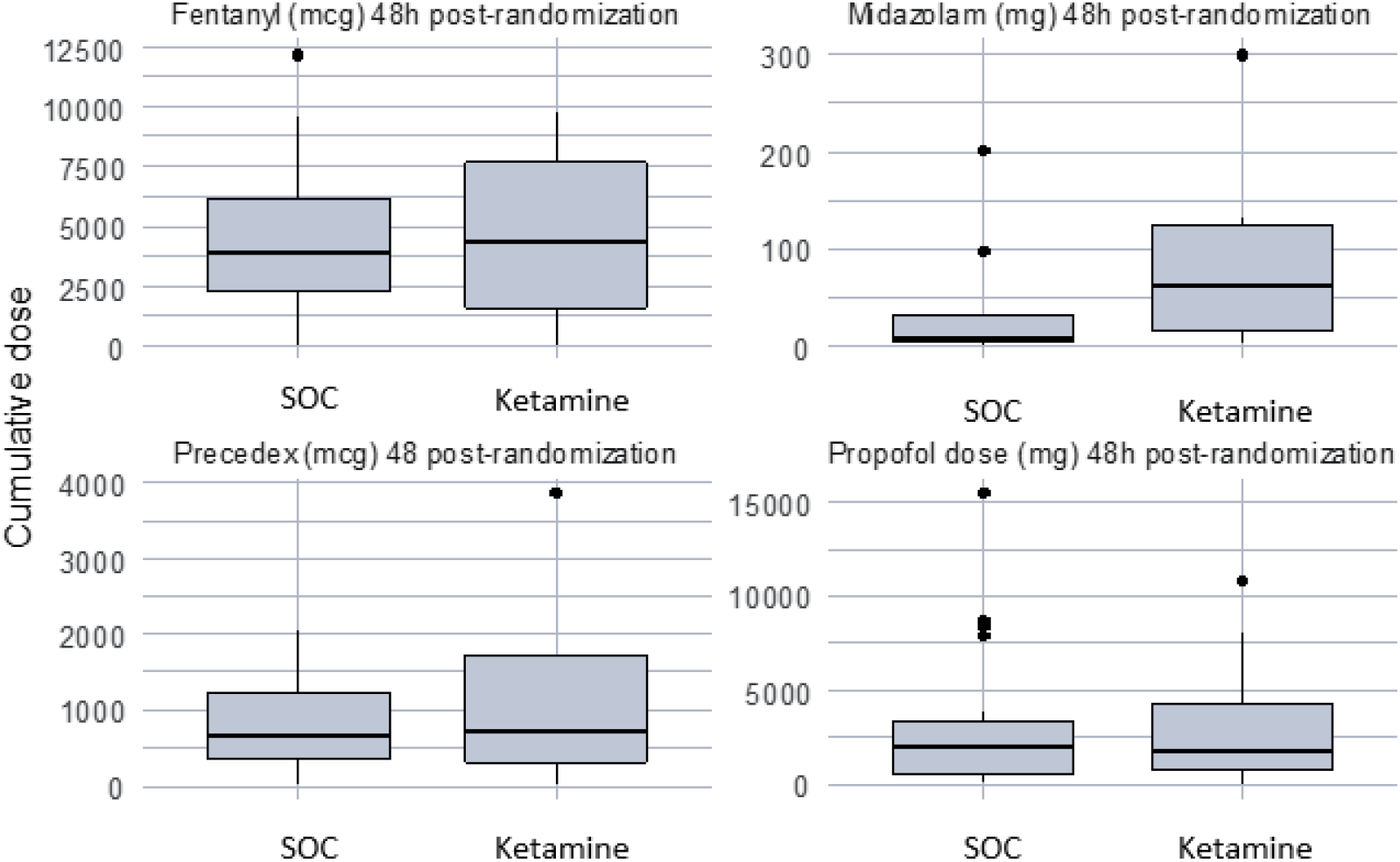
Box plots for cumulative doses of sedatives at 48 hours’ post-randomization.

**Supplementary Figure 4:**
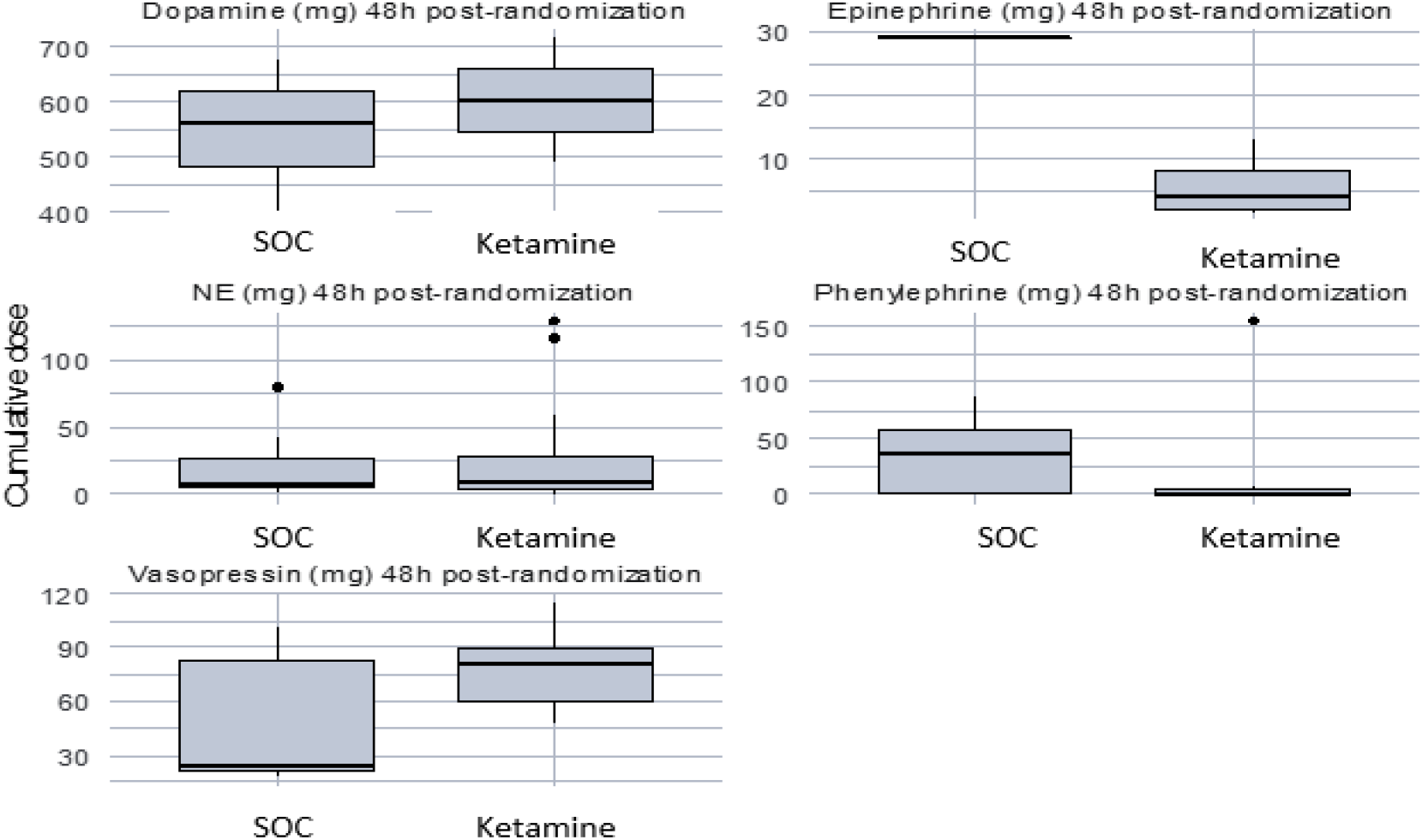
Box plots for cumulative doses of vasopressors at 48 hours’ post-randomization. SOC donates to standard of care

**Supplementary Table 7.** Sensitivity analysis for sedatives and vasopressors requirements excluding patients started on atracurium post-randomization.

|  | **SOC (N=41)** | **Ketamine (N=35)** | **P** |
| --- | --- | --- | --- |
| Patient on fentanyl within 48h, N (%) | 41 (100) | 34 (97.1) | 0.46 |
| Fentanyl route of administration (48h) , N (%) |  |  | 0.33 |
| Infusion | 35 (85.4) | 30 (88.2) |  |
| PRN bolus | 3 (7.32) | 0 (0) |  |
| Both | 3 (7.32) | 4 (11.8) |  |
| Cumulative dose of fentanyl (mcg) 48h post-randomization | 3817 (2200-5900) | 3400 (1500-6298) | 0.87 |
| Daily dose of fentanyl (mcg/Kg) 48h post-randomization | 66.1 (32.7-103) | 61.4 (19.5-108) | 0.91 |
| Patient on propofol 48h post-randomization, N (%) | 22 (53.7) | 20 (57.1) | 0.94 |
| Propofol route of administration (48h) , N (%) |  |  | 0.09 |
| Infusion | 22 (100) | 17 (85) |  |
| PRN bolus | 0 (0) | 1 (5) |  |
| Both | 0 (0) | 2 (10) |  |
| Cumulative propofol dose (mg) 48h post-randomization | 2161 (398-3406) | 1688 (778-4272) | 0.88 |
| Propofol daily dose (mg/kg) 48h post-randomization | 33.6 (7.65-58.5) | 27.8 (9.62-56.7) | 0.76 |
| Patient on midazolam within 48h post-randomization, N (%) | 7 (17.1) | 4 (11.4) | 0.71 |
| Midazolam route of administration (48h) , N (%) |  |  | 0.49 |
| Infusion | 3 (42.9) | 1 (25) |  |
| PRN bolus | 4 (57.1) | 2 (50) |  |
| Both | 0 (0) | 1 (25) |  |
| Cumulative dose of midazolam (mg) 48h post-randomization | 6 (4- 54.5) | 58.5 (12.5-110) | 0.29 |
| Midazolam daily dose (mg/kg) 48h post-randomization | 0.21 (0.09-0.72) | 0.74 (0.19-1.43) | 0.45 |
| Dexmedetomidine starter within 24h post-randomization, N (%) | 12 (29.3) | 4 (11.4) | 0.11 |
| Cumulative dose of Dexmedetomidine (mcg) 48 post-randomization | 667 (357-1222) | 711 (310-1730) | 0.90 |
| Daily dose of Dexmedetomidine (mcg/Kg) 48h post-randomization | 9.34 (5.33-22) | 18 (4.67- 35.5) | 0.63 |
| Patient on Norepinephrine within 48 h, N (%) | 25 (60.89) | 21 (60.0) | 1 |
| Cumulative dose of Norepinephrine (mg) 48h post-randomization | 8.07 (5.27 - 20.07) | 8.21 (3.66-28) | 0.97 |
| Patient of epinephrine within 48h post-randomization, N (%) | 1 (2.44) | 2 (5.71) | 0.59 |
| Cumulative dose of epinephrine (mg) 48h post-randomization | 29.2 (29.2- 29.2) | 3.805 (1.52 - 6.09) | 0.22 |
| Patient on phenylephrine within 48h post-randomization, N (%) | 5 (12.2) | 5 (14.29) | 1 |
| Cumulative dose of phenylephrine (mg) 48h post-randomization | 36 (0.3-72.55) | 0.6 (0.213-81.3) | 0.75 |
| Patient on dopamine within 48h post-randomization, N (%) | 3 (7.32) | 2 (5.71) | 1 |
| Cumulative dose of dopamine (mg) 48h post-randomization | 562.56 (401.74 - 675.84) | 602.3 (489.6- 715) | 0.56 |
| Patient on vasopressin within 48h post-randomization, N (%) | 7 (17.07) | 5 (14.29) | 1 |
| Cumulative dose of vasopressin (units) 48h post-randomization | 48 (21.6 - 95.57) | 69.6 (49.2-104.4) | 0.41 |

Data presented as n (%), mean ± sd, or median (interquartile range). SOC donates to standard of care

## Notes

**Conflicts of Interest and Source of Funding:** None declared

### Competing Interest Statement

The authors have declared no competing interest.

### Clinical Trial

NCT04075006

### Funding Statement

No funding. This trial was investigator-initiated and all study authors are employees at King Faisal Specialist Hospital and Research Center (KFSH&RC), which has not provided any research grant for this particular project. All authors volunteered their time and used local resources to conduct the study.

### Author Declarations

The study was conducted in accordance with the principles of the Declaration of Helsinki. Our institution [King Faisal Specialist Hospital and Research Center (KFSH&RC)] research ethics committee (REC) and office of research affair approved the study with the Research Advisory Council (RAC) number 2191 187. Given the need to enroll patients in an expedited manner within the 24-hour window, verbal consent from a guardian/next of kin was allowed and documented in the chart. Written consent was obtained as soon as the next of kin became available. During the COVID-19 pandemic, where patients family visits were prohibited, we were unable to obtain written consent. Therefore, we granted approval from our research ethics committee for the following consenting process: verbal consent from the family over the phone or a conference call to explain the nature of the study, as detailed in the written informed consent. A witness who was not part of this research study was present during the verbal consenting process and documentation was completed in the patient chart.

### Summary of Updates

we made some revision to clarify that it is pilot feasibility trial

